# Variation in US Drug Overdose Mortality Within and Between Hispanic/Latine Subgroups: A Disaggregation of National Data

**DOI:** 10.1101/2022.01.10.22268859

**Authors:** Manuel Cano, Camila Gelpí-Acosta

## Abstract

This study examined differences across Latine heritage groups (i.e., Mexican, Puerto Rican, Cuban, Dominican, Central American, South American) in rates of US drug overdose mortality. The study utilized 2015-2019 mortality data from the National Center for Health Statistics for 29,137 Hispanic individuals who died of drug overdose. Using population estimates from the American Community Survey, age-standardized drug overdose mortality rates were calculated by specific Latine heritage and sex, nativity, educational attainment, and geographic region. Standardized rate ratios (SRRs), incidence rate ratios (IRRs) from negative binomial regression models, and 95% Confidence Intervals (CIs) were calculated, and multiple imputation was used for missing Latine heritage group in select models.

Drug overdose mortality rates in the Puerto Rican heritage population were more than three times as high as in the Mexican heritage population (IRR 3.61 [95% CI 3.02-4.30] in unadjusted model; IRR 3.70 [95% CI 3.31-4.15] in model adjusting for age, sex, nativity, educational attainment, and region; SRR 3.23 [95% CI, 3.15-3.32] in age-standardized model with missing Hispanic heritage imputed). Higher age-standardized rates of drug overdose mortality were observed in males than females across all Latine groups, yet the magnitude of the sex differential varied by Latine heritage. The relationship between drug overdose mortality and nativity differed by Latine heritage; in all groups except Puerto Rican, overdose mortality rates were significantly higher in the US-born than those not US-born. In contrast, overdose mortality rates were significantly lower in US-born Puerto Ricans than in Puerto Ricans who were not US-born (e.g., born in Puerto Rico; SRR, 0.84 [95% CI 0.80-0.88]). The relationship between drug overdose mortality and educational attainment (for ages 25+) also varied between Latine groups.

The diverse subgroups comprising the US Latine population vary not only in rates of drug overdose mortality, but also in demographic risk factors for fatal drug overdose.

## 1. Introduction

Drug overdoses claimed more than 100,000 lives in the United States (US) in the year-long period ending April 2021 [1]. Although the national drug overdose mortality rate for Hispanics is lower than Non-Hispanic (NH) White or Black populations [2], overdose mortality is increasing among Hispanics [3–4], and rates of overdose mortality in Hispanics exceed rates in other racial/ethnic groups in certain states and/or cities [5–6]. The linguistic and cultural diversity within the Hispanic/Latine population, as well as the structural barriers faced by many Hispanics/Latines, warrant in-depth examination of drug overdose mortality within this diverse population in order to inform interventions that equitably address risk of overdose mortality.

Although numerous substance use-related outcomes have been well-researched in Latine populations [7–13], relatively less is known about the most recent patterns of drug overdose mortality in Latine subgroups. The present study builds on prior literature to address several gaps in our understanding of drug overdose mortality in Latines in the midst of the contemporary US overdose crisis. Although constrained by the categories available in national data and the limitations of classifying individuals into groups, we aim to apply LatCrit concepts of multidimensionality and anti-essentialism [14–17] to US mortality data from 2015-2019. In consideration of *multidimensionality*, we examine several dimensions of identities (e.g., specific heritage, nativity, educational attainment) and how these identities shape risk. With respect to *anti-essentialism*, we acknowledge the inherent heterogeneity in the Hispanic/Latine population, rather than considering Hispanics a monolithic ethnic group.

LatCrit theorists note the Eurocentric hierarchy implicit in the order of the terms listed in the US ethnic category “Spanish-Hispanic-Latina/o” (i.e., with Spanish listed first [18]). In the present study, we use the term “Hispanic” when referring to the category available in the study’s data source, which includes individuals identified as Spaniard. Meanwhile, we use the term “Latine” (a gender-inclusive version of Latina/o, comparable to “Latinx”) to refer to individuals who trace their heritage to Latin America, as many individuals often categorized as “Hispanic” have diverse ancestries and do not necessarily speak Spanish or identify with Spanish ancestry.

## 2. Background and Research Questions

### 2.1 Overdose Mortality Differences by Sex, Nativity, and Educational Attainment

In an analysis of drug overdose deaths in the US in 2017, the demographic characteristics of Latine decedents varied across heritage groups (e.g., Mexican, Cuban, Puerto Rican) [3]. At the same time, it is not clear to what extent these heritage-based variations in the demographics of overdose *decedents* could be attributed to heritage-based variation in the demographics of their underlying *populations*. The population distributions of different Latine subgroups vary by several characteristics. For example, an estimated 46.6% of the Dominican-heritage US population identifies as male, compared to 51.2% male in the Central American-heritage US population [19]. Variation by place of birth is even more pronounced, as only 33% of the Mexican-heritage population in the US is foreign-born, in contrast to 63% of the South American-heritage population in the US [20]. Finally, the distribution of educational attainment also varies by Latine heritage group, with 13.4% of the Mexican-heritage US population (aged 25 or older) reporting obtaining a Bachelor’s degree or higher, compared to 38.6% of the South American-heritage population [19].

Prior research has identified sex, nativity, and educational attainment as sources of variation in rates of drug overdose mortality in the general population and in large racial/ethnic categories [21–23]. First, with respect to sex differences, age-standardized drug overdose mortality rates are more than twice as high for men as women in the US population overall, and this sex differential is more pronounced in NH Black or Hispanic populations than the NH White population [23]. In the Puerto Rican US population specifically, age-standardized rates of drug overdose mortality in 2018 were more than three times as high in men as women [24]. Nonetheless, little is known about sex differentials in overdose mortality for other Latine heritage groups (e.g., Mexican, Cuban). Second, with respect to nativity or place of birth, drug overdose mortality rates are notably higher in those born in US states/territories/DC, relative to the foreign-born, and this nativity differential is more pronounced for NH Black, Hispanic, and NH Asian/Pacific Islander populations than NH Whites [21]. For Puerto Rican-heritage individuals, age-standardized rates of drug overdose mortality are higher in island-born, rather than US state-born, men [25]. However, it is unclear how nativity shapes risk of overdose mortality for other Latine heritage subgroups. Third, with respect to educational attainment, age-standardized drug overdose mortality rates are highest among those with the lowest level of education, both in the US population overall and in the NH White population [22]. Nonetheless, it is unclear to what extent this pattern is observed across specific Latine heritage groups.

The proceeding gaps in the literature inform the present study’s first research question (RQ): RQ1. What is the relationship between drug overdose mortality and sex, nativity, and educational attainment within each Latine heritage group?

### 2.2 Overdose Mortality Differences by Heritage and Geographic Region

Prior research has documented substantial variation in national rates of drug overdose mortality across Latine heritage groups, with the highest rates of overdose mortality observed in the Puerto Rican heritage group, followed by Dominican and Cuban heritage, next followed by Mexican heritage, and finally followed by Central or South American heritage [3]. Nonetheless, it is unclear whether this national pattern is observed across all US geographic regions. Risk of fatal overdose varies widely by region of the US [26], in part due to differences in regional drug supplies. For example, in the Northeast, heroin supplies are dominated by illicitly-manufactured fentanyl, which dramatically increases risk of a fatal overdose [27–28]. This geographic variation in risk of overdose mortality leads to the study’s second research question:

*RQ2: How do rates of drug overdose mortality by Latine heritage group compare across the four US geographic regions?*

Latine heritage groups are concentrated in different regions of the US; for example, Puerto Ricans comprise the largest Latine group in the Northeast, while Mexicans represent the largest Latine group in the other three US regions [19]. Mexicans account for 78% of the Hispanic population in the Los Angeles-Long Beach, California area, yet they represent only 3% of the Hispanic population in the Miami-Hialeah, Florida area, where Cubans account for 54% of Hispanics [29]. Considering the different regions where Latine heritage groups reside, as well as the geographic variation in the risk environment for drug overdose, it is unclear whether heritage differences in drug overdose mortality rates would be observed *even* after accounting for geographic region of residence and other demographic characteristics that vary between subgroups. Therefore, the study’s third research question asks: *RQ3. Do Latine heritage differences in drug overdose mortality rates persist after adjusting for geographic region of residence, nativity, educational attainment, sex, and age?*

### 2.3 Missing Specific Hispanic Heritage

Since demographics on the death certificate are furnished by a funeral director, race/ethnicity information is sometimes misclassified in mortality data [30]. Instances of misclassification include decedents who were categorized under a race/ethnicity group that did not match the way these decedents would have self-identified during life, as well as Hispanic decedents for whom the information provided on the death certificate was insufficient to identify a specific Hispanic heritage group. In an analysis of race/ethnicity death certificate identification from 1999-2011, the extent of misclassification (as assessed by race/ethnicity that did not match self-reported race/ethnicity from the Current Population Survey) varied by Hispanic subgroup, nativity, residence region, and coethnic concentration [30]. Misclassification was more notable for the Central and South American and “other Hispanic” groups relative to Mexican, Puerto Rican, or Cuban heritage [30]. In a 2017 analysis of drug overdose deaths in US Hispanics, 17.6% of the decedents were identified as “other Hispanic,” including both those identified as Spaniard as well as those with insufficient heritage information [3]. The considerable level of missingness of specific Hispanic heritage group information on the death certificate informs the present study’s last research question: *RQ4. Do Latine heritage differences in drug overdose mortality rates persist after imputing missing Hispanic heritage?*

## 3. Methods

### 3.1 Data Sources and Measures

Death certificate data were obtained via restricted-access Multiple Cause of Death files from the National Center for Health Statistics [31]. Demographic data on death certificates are generally supplied by a funeral director (often as informed by a next of kin [30]), while death information is specified by a medical certifier (e.g., coroner or medical examiner [32]). The present study included 29,137 deaths which met the following criteria: a) the death occurred in the 50 United States or DC between 2015-2019; b) the decedent was a US resident identified as of Hispanic heritage in the mortality data; c) the recorded “underlying cause of death” was drug poisoning of any intent (ICD-10 codes X40-44, X60-64, X85, or Y10-14); and d) the decedent’s age was not missing (three deaths excluded). In the 2015-2019 mortality data, 0.67% of drug overdose deaths lacked information on whether the decedent was of Hispanic origin; these decedents were not included.

Primary measures of interest included:

- Heritage, classified into eight mutually-exclusive categories based on Hispanic origin data from the death certificate: 1) Mexican; 2) Puerto Rican; 3) Cuban; 4) Dominican; 5) Central American; 6) South American; 7) Spaniard; and 8) specific Hispanic heritage missing/unclear [this category is detailed in the Appendix/Supplementary Material].
- Nativity/place of birth, classified into either US-born (born in the 50 United States or District of Columbia) or not US-born (born in a US territory or a foreign country)
- Sex (male or female, as recorded on the death certificate via observation or medical records [33])
- Age at time of death (in years, in categories <1, 1-4, 5-14, 15-24, 25-34, 35-54, 55-64, 65-74, 75-84, 85+)
- Educational attainment, classified into 8^th^ grade or lower, 9-12^th^ grade with no diploma, high school or GED, more than high school, or unknown. In the present study, educational attainment was examined only for individuals ages 25 or older.
- US Census Region (Northeast, South, Midwest, or West), based on the region corresponding to the state of residence of the decedent

In addition to the mortality data, the study used subpopulation estimates drawn from the Integrated Public Use Microdata Series’ (IPUMS) [34] representative samples of the American Community Survey 2015-2019 five-year estimates from the US Census Bureau. The Institutional Review Board of the University of Texas at San Antonio waived review of the present study’s analysis of the de-identified decedent data.

### 3.2 Statistical Analyses

Counts of drug overdose deaths from the NCHS data, as well as population estimates from the IPUMS, were tabulated by heritage, nativity, sex, age, educational attainment, and region of residence. Heritage categories used in these analyses included Mexican, Puerto Rican, Cuban, Dominican, Central American, and South American; the Spaniard category was not included due to small numbers of deaths.

#### 3.2.1 Calculation of Age-Adjusted Rates by Heritage and Key Demographics

Age-standardized drug overdose mortality rates were calculated using the direct method of age adjustment to the 2000 US standard population [35], with 95% Confidence Intervals calculated based on the gamma distribution, using the user-written program *distrate* [36]. Age-adjusted rates were calculated and plotted by heritage and sex, heritage and educational attainment (for ages 25+ only), heritage and nativity, and heritage and geographic region. Standardized rate ratios and 95% confidence intervals were used in planned comparisons of rates, while z-tests (alpha=0.05, two-tailed) were used for any post-hoc tests of significance of differences between rates.

#### 3.2.2 Regression of Drug Overdose Mortality on Heritage

In order to determine whether heritage differences in drug overdose mortality rates persist after accounting for heritage-based variation in region of residence, educational attainment, nativity, sex, and age, negative binomial regression was used with an offset term for the natural log of the subpopulation size. The unadjusted model included heritage group as the only predictor of overdose deaths, while the fully-adjusted model included age at time of death (in ten-year intervals), sex, nativity, US region of residence, and educational attainment. Both models were restricted to ages 25 and older due to the inclusion of educational attainment in the fully-adjusted model.

#### 3.2.3 Examination of Missingness of Hispanic Heritage Group

The ethnicity information for 17.48% of the 29,137 Hispanic drug overdose decedents lacked sufficient detail to classify these decedents into one of the seven Hispanic heritage groups (Mexican, Puerto Rican, Cuban, Dominican, Central American, South American, or Spain). The ethnicity codes which lacked sufficient heritage information are detailed in the Appendix, including “Hispanic” (with no further information on heritage) and “Latin American” (which may refer to any of multiple heritage groups). Decedents with such codes were classified as “Hispanic heritage group missing.” Proportions of drug overdose deaths with Hispanic heritage group missing were examined by demographic characteristics (sex, age category, educational attainment, marital status, and nativity), type of drug overdose (by intent and drugs involved), and state of residence.

#### 3.2.4 Imputation Procedures for Hispanic Heritage Group Missingness

The associations between missingness of Hispanic heritage group and observed variables in the data supported the assumption of Missing At Random (MAR), rather than Missing Completely At Random (MCAR) [37], although the possibility of Missing Not At Random cannot be systematically discounted. The Appendix provides detailed information about the assumptions of MAR for Hispanic missingness in the study. A polytomous logistic regression imputation model was utilized for multiple imputation. Predictors included decedent demographics and drugs involved in the overdose, as well as the county or state-level percentages of Latines identified as Mexican, Puerto Rican, Cuban, Dominican, Central American, or South American, respectively (using county-level data from the IPUMS American Community Survey five-year 2015-2019 estimates for all counties available and state-level data for counties where Latine heritage counts were unavailable due to population sizes). Details of all variables, as well as their association with missingness, are provided in the Appendix (Supplemental Tables 2 and 3). A total of 20 imputed datasets were generated, and subsequent analyses were conducted with the *mi estimate* suite of commands in Stata/MP 16.1. We used the imputed datasets to tabulate counts of drug overdose deaths by heritage group in order to estimate drug overdose mortality rates with missing Hispanic heritage imputed, standardizing rates to the 2000 US standard population.

##### 3.2.4.1 Evaluation of Performance of Imputation Model

The method used to evaluate the predictive accuracy of the imputation model was based on approaches used by Boslett and colleagues [38] and Montealegre and colleagues [39]:

- First, all decedents missing a specific Hispanic heritage group were dropped from the dataset in order to leave a “test” dataset with only complete responses for Hispanic heritage group, which we refer to as the “heritage” variable.
- In the “test” dataset, we created a duplicate variable for specific Hispanic heritage group, “heritage2,” and then randomly designated “heritage2” as missing for approximately 20% of the decedents in this test dataset.
- Next, we used the multiple imputation model (as specified in the preceding paragraph, with 20 imputed sets) in the “test” dataset to impute “heritage2” for those that had been randomly assigned as missing on this variable.
- We then calculated the proportion of decedents with an identical response for the variable “heritage” (that is, the heritage group that was actually recorded from the death certificate) and the variable “heritage2” (imputed heritage for those randomly assigned as missing), across the 20 imputed datasets, for decedents who had been randomly assigned as missing on “heritage2.”
- Finally, we used the *mi estimate* command, which uses pooled estimates across the multiple imputed datasets, to calculate what proportion each Hispanic heritage group represented in the imputed test dataset (that is, what proportion of responses on “heritage2” were Mexican, Puerto Rican, etc.). These proportions were referenced against the proportions of each group in the “heritage” variable (the variable for the heritage group actually recorded from the death certificate) in the test dataset.

## 4. Results

### 4.1 Drug Overdose Mortality Rates by Heritage and Sex, Education, and Nativity (RQ1)

Table 1 and Figure 1 provide age-standardized drug overdose mortality rates disaggregated by heritage and sex, heritage and nativity, and heritage and educational attainment. Table 1 also includes Standardized Rate Ratios (SRRs) with accompanying 95% CIs to compare rates by sex, nativity, and educational attainment across each heritage group. These results are based on observed data for complete cases only (i.e., without imputing missing Hispanic heritage), in contrast to results based on imputed data, which were calculated subsequently.

**Table 1.** Age-standardized drug overdose mortality rates (per 100,000) by Latine heritage and sex, nativity, and educational attainment, with Standardized Rate Ratios (SRRs) and accompanying 95% CIs, for US deaths 2015-2019

|  | Mexican | Puerto Rican | Cuban | Dominican | Central American | South American |
| --- | --- | --- | --- | --- | --- | --- |
| **Sex** |  |  |  |  |  |  |
| Male | 10.88 (10.65-11.10) | 44.27 (43.12-45.45) | 12.15 (11.27-13.07) | 17.94 (16.69-19.25) | 6.21 (5.82-6.64) | 6.78 (6.25-7.33) |
| Female | 3.85 (3.72-3.99) | 11.05 (10.49-11.63) | 4.73 (4.17-5.35) | 3.49 (3.00-4.04) | 1.41 (1.21-1.63) | 2.35 (2.06-2.68) |
| SRR, Male vs. Female | **2.82 (2.71-2.94)** | **4.01 (3.78-4.25)** | **2.57 (2.22-2.97)** | **5.14 (4.36-6.07)** | **4.40 (3.75-5.19)** | **2.88 (2.47-3.36)** |
| **Nativity** |  |  |  |  |  |  |
| US-Born | 12.13 (11.87-12.38) | 25.28 (24.48-26.11) | 13.70 (12.52-14.98) | 12.03 (10.32-14.16) | 7.54 (6.44-8.81) | 8.78 (7.75-9.96) |
| Not US-Born | 3.54 (3.39-3.70) | 30.25 (29.18-31.37) | 4.86 (4.35-5.57) | 9.37 (8.66-10.19) | 3.47 (3.22-3.75) | 3.35 (3.04-3.79) |
| SRR, US-Born vs. Not US-Born | **3.43 (3.26-3.60)** | **0.84 (0.80-0.88)** | **2.82 (2.39-3.25)** | **1.28 (1.08-1.54)** | **2.18 (1.82-2.59)** | **2.62 (2.19-3.08)** |
| **Educational Attainment<sup>a</sup>** |  |  |  |  |  |  |
| 8th grade or less | 6.19 (5.82-6.56) | 81.69 (75.41-88.29) | 14.67 (10.10-20.35) | 21.99 (17.85-26.68) | 3.92 (3.39-4.52) | 2.62 (1.48-4.21) |
| 9th-12th grade | 15.31 (14.70-15.95) | 79.59 (75.56-83.78) | 18.48 (15.08-22.35) | 17.80 (14.82-21.27) | 6.23 (5.30-7.36) | 10.72 (8.14-13.80) |
| HS/GED | 15.01 (14.56-15.48) | 65.18 (62.95-67.48) | 17.87 (16.16-19.71) | 24.80 (22.45-27.37) | 7.00 (6.26-7.84) | 10.85 (9.65-12.15) |
| >HS | 6.52 (6.24-6.82) | 12.84 (12.08-13.65) | 6.72 (5.95-7.56) | 5.98 (5.08-7.13) | 3.72 (3.22-4.32) | 3.78 (3.37-4.25) |
| SRR, 8th Grade or Less vs. >HS | 0.95 (0.88-1.02) | **6.36 (5.75-7.02)** | **2.18 (1.47-3.10)** | **3.68 (2.78-4.74)** | 1.05 (0.85-1.29) | 0.69 (0.39-1.13) |

*Notes.* <sup>a</sup>Educational attainment only includes those age 25 or older at the time of death. Bold indicates that SRR is statistically significantly different from 1.00 at p<0.05. "Not US-born" includes both those born in a foreign country and those born in a US territory such as Puerto Rico.
*Abbreviations.* CI, Confidence Interval; HS, High School; GED, General Education Development

**Figure 1.**
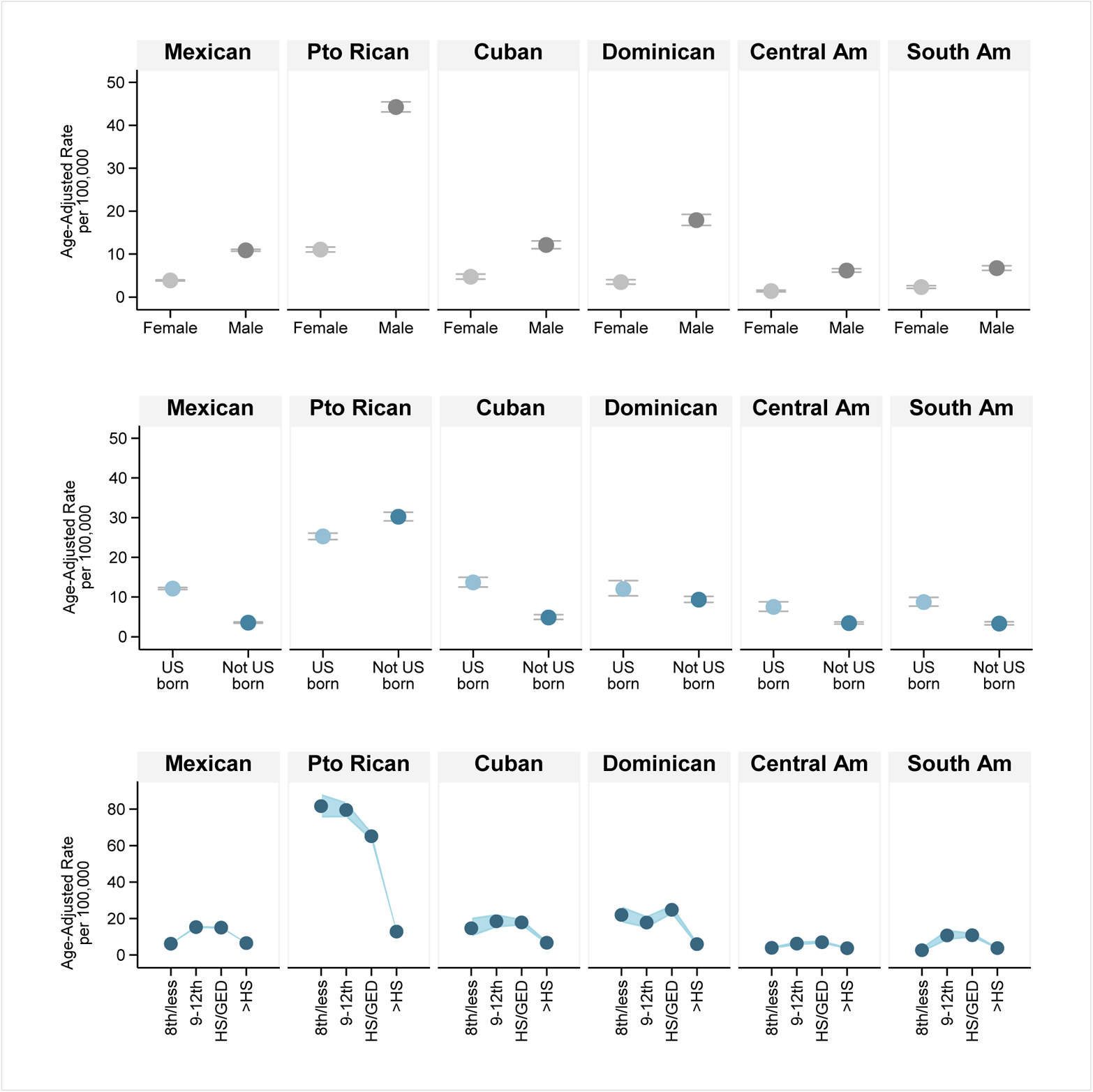
Age-standardized drug overdose mortality rates for US Latines (2015-2019), by heritage and: (a) sex; (b) nativity; and (c) educational attainment^a^. *Notes.* ^a^Educational attainment only includes those age 25 or older at the time of death. “Not US-born” includes both those born in a foreign country and those born in a US territory such as Puerto Rico. *Abbreviations.* Pto, Puerto; Am, American; HS, High School; GED, General Education Development

#### 4.1.1 Sex

Age-standardized drug overdose mortality rates were higher in males than females across all heritage groups examined, although the magnitude of the sex gap differed by heritage. The largest *absolute difference* in rates between males and females (rate in males minus rate in females) was observed among Puerto Ricans (44.27 per 100,000 in males versus 11.05 in females), while the largest *relative difference* in rates by sex (rate in males divided by rate in females) was observed among Dominicans, with drug overdose mortality rates more than five times as high in Dominican males as Dominican females (SRR 5.14 [95% CI, 4.36-6.07]).

#### 4.1.2 Nativity

Puerto Rican heritage was the only heritage group in which age-standardized drug overdose mortality rates were *lower* for the US-born than those not born in the US (SRR 0.84 [95% CI, 0.80-0.88]). In all other Latine heritage groups, drug overdose mortality rates were higher among the US-born than those not US-born. The relative nativity differential was largest for the Mexican heritage group, with rates of overdose mortality more than three times as high in the US-born as the foreign-born (SRR 3.43 [95% CI, 3.26-3.60]).

#### 4.1.3 Educational Attainment

The association between drug overdose mortality and educational attainment (for ages 25+) varied by heritage. In the Mexican, Central American, and South American groups, age-standardized rates of drug overdose mortality peaked among those with a 9-12^th^ grade education or high school diploma/GED, while rates were comparably lower for those with the lowest (8^th^ grade or less) and highest (more than high school) educational levels. In contrast, in the Puerto Rican, Cuban, and Dominican groups, drug overdose mortality rates were significantly lower for those with the highest education (more than high school) relative to the lowest education (8^th^ grade or less). The largest educational differential was observed in Puerto Rican heritage, with drug overdose mortality rates more than six times as high in those with the lowest relative to the highest educational level (SRR 6.36 [95% CI, 5.75-7.02]).

### 4.2 Drug Overdose Mortality Rates by Heritage Across US Regions (RQ2)

As presented in Figure 2, age-standardized drug overdose mortality rates were significantly higher in the Puerto Rican group than in any other heritage group across all US regions, except the West, where the difference between Puerto Rican and Dominican rates was not statistically significant. Drug overdose mortality rates were highest in the Northeast region for all heritage groups except Mexican; for the Mexican heritage group, rates were highest in the Midwest.

**Figure 2.**
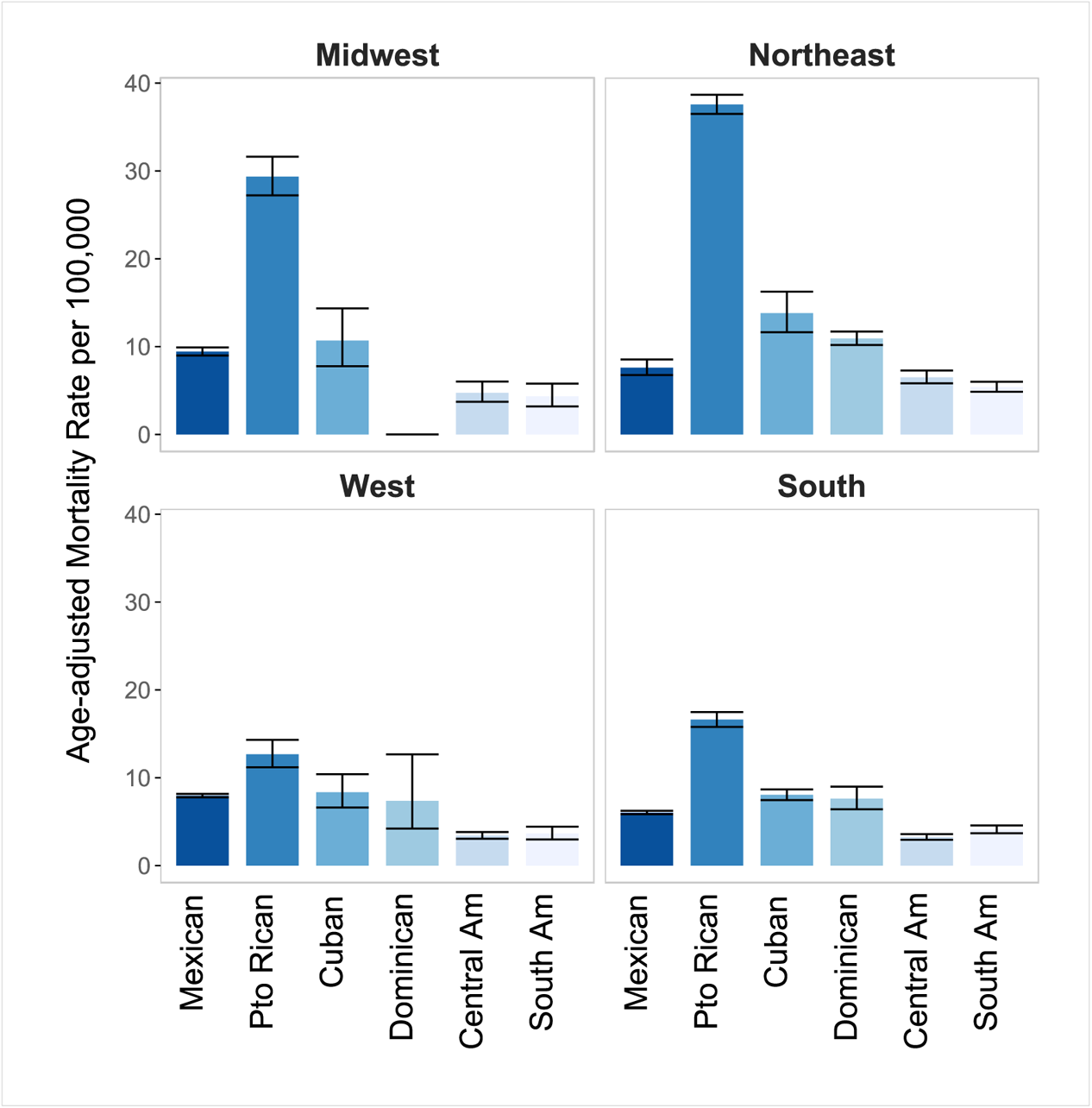
Age-standardized drug overdose mortality rates and 95% CIs for US Latines (2015-2019) by US Census Region of residence. Abbreviations. CI, Confidence Interval; Pto, Puerto; Am, American Notes. Rate for Dominican heritage in the Midwest was suppressed due to small numbers.

### 4.3 Heritage Differences in Unadjusted and Adjusted Models (RQ3)

Table 2 provides Incidence Rate Ratios (IRRs) and accompanying 95% CIs for the unadjusted regression model (regressing overdose deaths on heritage group) and the fully-adjusted model (adjusted for age, sex, nativity, US region of residence, and educational attainment). These models were limited to ages 25 and older, since educational attainment is a measure less relevant in younger ages. In both the unadjusted and fully-adjusted model, rates of drug overdose mortality were more than three times as high in Puerto Rican as Mexican heritage (IRR 3.61 [95% CI, 3.02-4.30] in the unadjusted model; IRR 3.70 [95% CI, 3.31-4.15] in the fully-adjusted model). Across both models, drug overdose mortality rates were significantly higher in Cuban or Dominican heritage relative to Mexican heritage, while significantly lower in Central or South American than Mexican heritage.

**Table 2.** Results from the negative binomial regression of drug overdose deaths on Latine heritage among individuals age 25 or older at the time of death, 2015-2019.

|  | Unadjusted Model<sup>a</sup> |  | Fully Adjusted Model<sup>b</sup> |  |
| --- | --- | --- | --- | --- |
|  | IRR (95% CI) | p value | IRR (95% CI) | p value |
| Hispanic heritage |  |  |  |  |
| Mexican | 1.00 (reference) | - | 1.00 (reference) | - |
| Puerto Rican | 3.61 (3.02-4.30) | <0.001 | 3.70 (3.31-4.15) | <0.001 |
| Cuban | 1.40 (1.13-1.73) | 0.002 | 1.61 (1.39-1.86) | <0.001 |
| Dominican | 1.27 (1.01-1.59) | 0.042 | 1.35 (1.15-1.59) | <0.001 |
| Central American | 0.63 (0.51-0.77) | <0.001 | 0.65 (0.57-0.75) | <0.001 |
| South American | 0.72 (0.58-0.89) | 0.002 | 0.82 (0.71-0.95) | 0.009 |
| Information indices<sup>c</sup> |  |  |  |  |
| df | 7 |  | 21 |  |
| AIC | 9442.66 |  | 7813.12 |  |
| BIC | 9483.94 |  | 7936.94 |  |

Notes. <sup>a</sup>Regression of deaths on Latine heritage, with offset term for sub-population size; <sup>b</sup>Regression of deaths on Latine heritage, with offset term for population size, adjusted for age, sex, nativity, US Census region, and educational attainment; <sup>c</sup>df, degrees of freedom; AIC, Akaike's information criterion; BIC, Bayesian information criterion. Abbreviations: IRR, incidence-rate ratio; CI, confidence interval.

### 4.4 Missingness of Specific Hispanic Heritage

Table 3 provides the proportion of Hispanic drug overdose decedents missing specific Hispanic heritage, by characteristics of the decedent or the death. The largest proportions of missingness of specific Hispanic heritage were observed among decedents who were also missing other death certificate demographics (i.e., those with a response of “unknown” for educational attainment or marital status). Differences in missingness by nativity were also observed, as 22.5% of those born in the 50 states/DC were missing a specific Hispanic heritage, compared to only 8.1% of the foreign-born and 0.1% territory-born. Figure 3 depicts proportions of missingness of specific Hispanic heritage in each state; states with percentages based on fewer than 10 decedents were excluded due to NCHS confidentiality requirements. The proportion of missingness for specific Hispanic heritage was highest in New Mexico (86.5% of Hispanic overdose decedents were missing specific Hispanic heritage in New Mexico), followed by Colorado (63.5% missing).

**Table 3.**
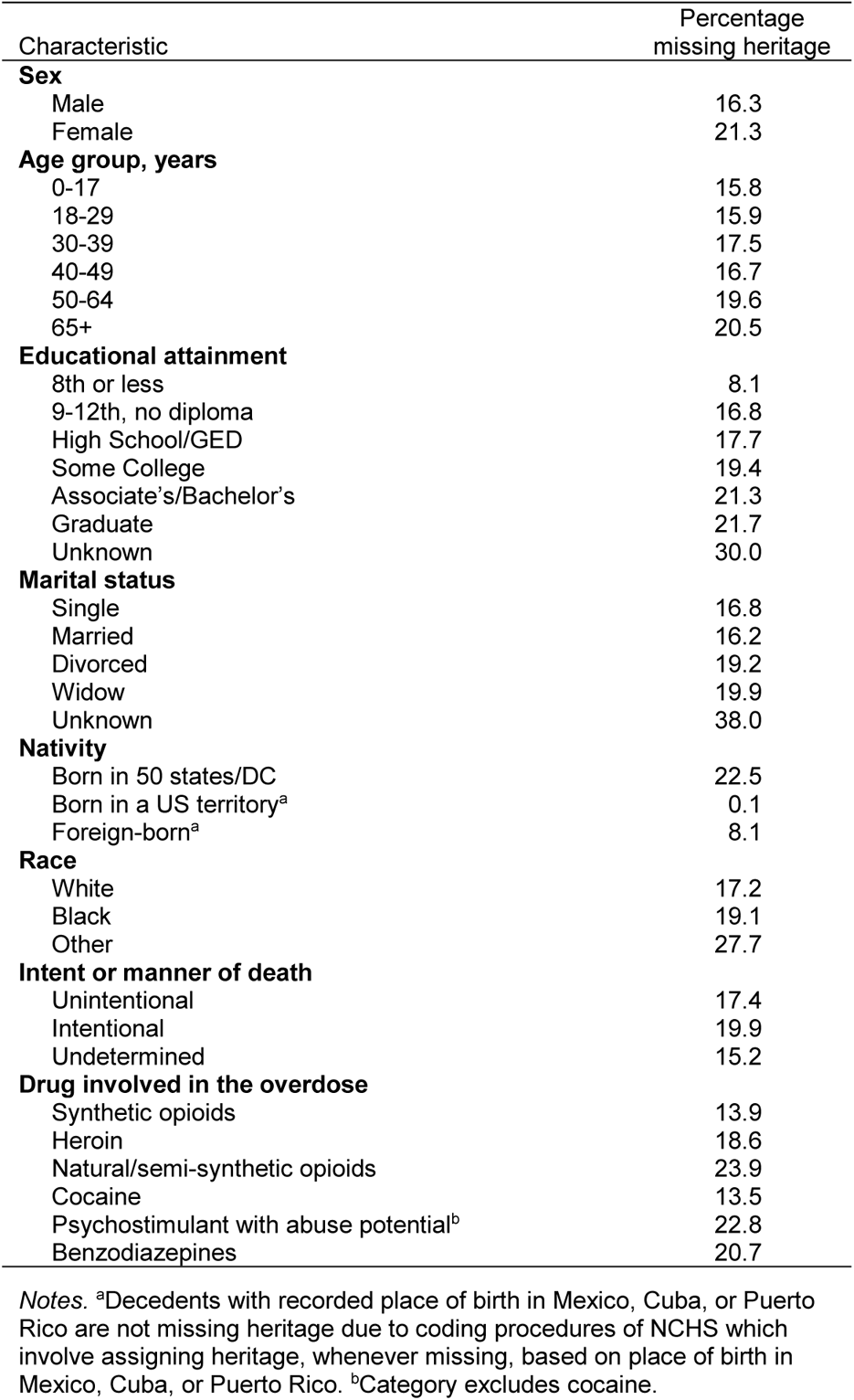
Percentage of missingness of specific Hispanic heritage, by selected characteristics, for drug overdose deaths among individuals identified as of Hispanic heritage, 2015-2019

| Characteristic | Percentage missing heritage |
| --- | --- |
| **Sex** |  |
| Male | 16.3 |
| Female | 21.3 |
| **Age group, years** |  |
| 0-17 | 15.8 |
| 18-29 | 15.9 |
| 30-39 | 17.5 |
| 40-49 | 16.7 |
| 50-64 | 19.6 |
| 65+ | 20.5 |
| **Educational attainment** |  |
| 8th or less | 8.1 |
| 9-12th, no diploma | 16.8 |
| High School/GED | 17.7 |
| Some College | 19.4 |
| Associate's/Bachelor's | 21.3 |
| Graduate | 21.7 |
| Unknown | 30.0 |
| **Marital status** |  |
| Single | 16.8 |
| Married | 16.2 |
| Divorced | 19.2 |
| Widow | 19.9 |
| Unknown | 38.0 |
| **Nativity** |  |
| Born in 50 states/DC | 22.5 |
| Born in a US territory<sup>a</sup> | 0.1 |
| Foreign-born<sup>a</sup> | 8.1 |
| **Race** |  |
| White | 17.2 |
| Black | 19.1 |
| Other | 27.7 |
| **Intent or manner of death** |  |
| Unintentional | 17.4 |
| Intentional | 19.9 |
| Undetermined | 15.2 |
| **Drug involved in the overdose** |  |
| Synthetic opioids | 13.9 |
| Heroin | 18.6 |
| Natural/semi-synthetic opioids | 23.9 |
| Cocaine | 13.5 |
| Psychostimulant with abuse potential<sup>b</sup> | 22.8 |
| Benzodiazepines | 20.7 |

*Notes.* <sup>a</sup>Decedents with recorded place of birth in Mexico, Cuba, or Puerto Rico are not missing heritage due to coding procedures of NCHS which involve assigning heritage, whenever missing, based on place of birth in Mexico, Cuba, or Puerto Rico. <sup>b</sup>Category excludes cocaine.

**Figure 3.**
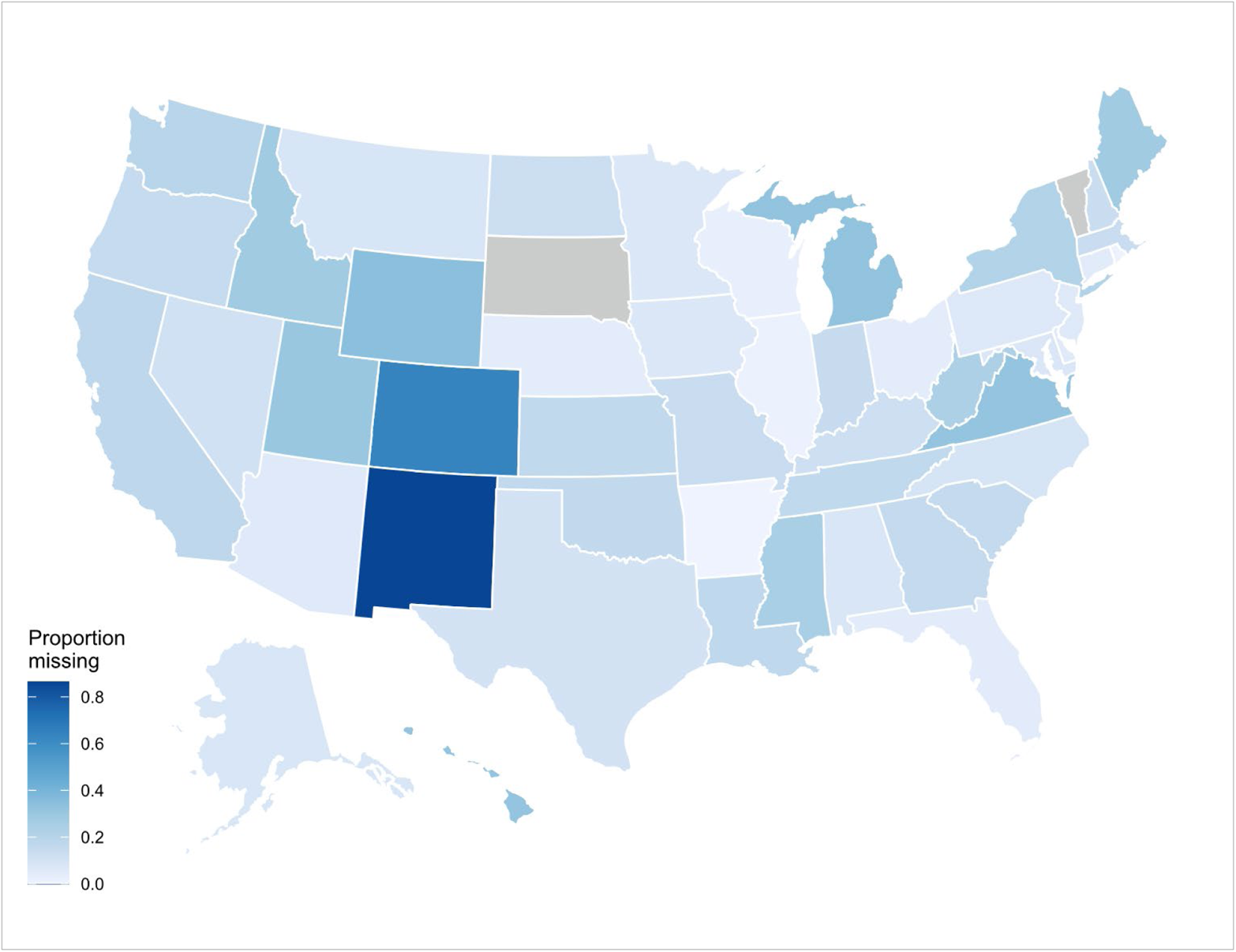
State-level proportion of missingness of specific Hispanic heritage group for Hispanic drug overdose deaths, 2015-2019. *Notes.* States with percentages based on fewer than 10 decedents are excluded due to NCHS confidentiality requirements.

### 4.5 Results after Multiple Imputation (RQ4)

Table 4 provides age-standardized drug overdose mortality rates and Standardized Rate Ratios by heritage group, using both the observed data (only including decedents with complete heritage information) and the imputed data (with missing heritage imputed). The age-standardized drug overdose mortality rates estimated with the imputed data were significantly higher than the observed (unimputed) mortality rates for each Latine heritage group.

SRRs (comparing age-standardized drug overdose mortality rates in each heritage group against the rate in Mexican heritage) were relatively similar in the observed (unimputed) and imputed data. In both observed and imputed data, estimated age-standardized rates of drug overdose mortality in the Puerto Rican heritage group were more than three times as high as the Mexican heritage group, although the difference was slightly less pronounced in the imputed data (SRR 3.23 [95% CI, 3.15-3.32]) than the observed data (SRR 3.68 [95% CI, 3.57-3.79]). In both observed and imputed data, age-standardized rates of drug overdose mortality were significantly higher in the Dominican heritage than Mexican heritage while significantly lower in the Central American or South American groups. In the observed data, rates were significantly higher in Cuban heritage than Mexican heritage, yet the difference between Cuban and Mexican heritage was not significantly different from zero in the imputed data.

**Table 4.** Age-standardized drug overdose mortality rates and Standardized Rate Ratios (SRRs) by Latine heritage, with observed or imputed data, 2015-2019

| Heritage | <u>Observed Data</u> |  | <u>Imputed Data</u> |  |
| --- | --- | --- | --- | --- |
|  | Rate per 100,000<br>(95%CI) | SRR (95% CI) | Rate per 100,000<br>(95%CI) | SRR (95% CI) |
| Mexican | 7.46 (7.32-7.59) | 1.00 (reference) | 9.58 (9.43-9.73)<sup>§</sup> | 1.00 (reference) |
| Puerto Rican | 27.45 (26.81-28.10) | **3.68 (3.57-3.79)** | 30.95 (30.27-31.63)<sup>§</sup> | **3.23 (3.15-3.32)** |
| Cuban | 8.61 (8.07-9.17) | **1.15 (1.08-1.23)** | 9.57 (9.01-10.16)<sup>§</sup> | 1.00 (0.94-1.06) |
| Dominican | 10.07 (9.45-10.73) | **1.35 (1.26-1.44)** | 11.76 (11.09-12.47)<sup>§</sup> | **1.23 (1.16-1.31)** |
| Central American | 3.94 (3.71-4.18) | **0.53 (0.50-0.56)** | 4.57 (4.32-4.83)<sup>§</sup> | **0.48 (0.45-0.51)** |
| South American | 4.49 (4.20-4.80) | **0.60 (0.56-0.65)** | 5.20 (4.88-5.53)<sup>§</sup> | **0.54 (0.51-0.58)** |

*Notes.* All rates are directly standardized to the 2000 US standard population. Observed data are for complete cases only; imputed data use imputed Hispanic heritage whenever missing. SRRs are bolded if their 95% CI does not include the value of 1.00. <sup>§</sup>Significantly different from rate in observed data, per z-test (two-tailed, alpha=0.05). *Abbreviations.* CI, Confidence Interval; SRR, Standardized Rate Ratio.

Finally, drug overdose mortality rates by specific Latine heritage were compared against rates observed in NH White and NH Black populations, as a frame of reference. Calculated using the same source of population estimates (ACS five-year estimates via IPUMS) and directly standardized to the same age distribution of the 2000 US standard population, the 2015-2019 drug overdose mortality rate was 25.55 (95% CI, 25.45-25.65) in the NH White population and 19.90 (95% CI, 19.70-20.09) in the NH Black population. In both the observed and imputed Hispanic heritage data, drug overdose mortality rates were significantly higher in the Puerto Rican heritage group (observed: 27.45 [95% CI, 26.81-28.10]; imputed: 30.95 [95% CI, 30.27-31.63]) than the NH White or NH Black populations, while significantly lower in all other Latine heritage groups than in NH White or NH Black populations.

#### 4.5.1 Performance of Imputation Model

In the test dataset with simulated missingness of heritage, the *imputed* Hispanic heritage group identically matched the *observed* Hispanic heritage group for 72.7% of all simulated missing records across 20 imputed datasets. Using the *mi estimate* command (which uses pooled estimates across the multiple imputed datasets), we calculated the proportional share each Hispanic heritage group represented when using the imputed heritage variable in the simulated missing heritage test dataset vs. the proportional shares observed in the heritage data on complete cases: Mexican 53.08% vs. 53.09%; Puerto Rican 29.85% vs 29.94%; Cuban 4.21% vs 4.17%; Dominican 4.14% vs. 4.14%; Central American 4.95% vs. 4.87%; South American 3.57% vs. 3.60%; and Spain 0.20% vs. 0.20%.

## 5. Discussion

The present study examined differences across Latine heritage groups (e.g., Mexican, Puerto Rican) in rates of drug overdose mortality, extending findings from prior literature by including both unadjusted and adjusted models, using both complete case analysis and imputing missing heritage group, and examining the intersection of heritage and several demographic characteristics. Aligned with LatCrit’s anti-essentialist foundation (i.e., Latines are not a monolithic group), findings emphasized the dramatic variation in rates of drug overdose mortality across Latine groups. Simply put, not all Latine populations are affected equally by drug overdose mortality, and hence the Hispanic concept obscures [16] the complexity of Latine populations and their varying health vulnerabilities. Moreover, Latine heritage groups also differ in terms of the relationship between drug overdose mortality and sex, nativity, and educational attainment, although the measures used in the present study are admittedly limited in their ability to capture complex social phenomena such as individual and collective histories, migration trajectories, colonialism, and cultural values, practices, and norms. LatCrit’s multidimensionality precept is not only relevant but also necessary to understand the health vulnerabilities of Latines in the US, with racism, nativism, sexism, and homophobia representing some of the multiple dimensions coalescing in the in the everyday experiences of different Latines in the US, [14] impacting health outcomes such as fatal overdose.

### 5.1 Differences in Drug Overdose Mortality Rates by Latine Heritage Group

A previous analysis of 2017 US data found that age-standardized rates of drug overdose mortality were significantly and substantially higher in the Puerto Rican heritage group than the Mexican, Cuban, Dominican, Central American, or South American groups [3]. Results of the present study add that this finding is also observed across multiple years (2015-2019 pooled), across three of the four geographic regions of the US (all regions except the West), and whether using observed data only (complete case analysis) or procedures for imputing missing Hispanic heritage group. Furthermore, the present study’s results indicate that Latine heritage group differences in drug overdose mortality persist *even after* adjusting for variation in key demographics such as geographic region of residence, nativity, educational attainment, sex, and age.

Overall, the study’s results across various analyses consistently support the disproportionately high rates of drug overdose mortality in the Puerto Rican heritage group. Nevertheless, reasons for heritage differences in overdose mortality are less clear. Results from the adjusted regression models suggest that the higher rates of drug overdose mortality in Puerto Ricans relative to other Latine groups cannot be primarily attributed to the geographic regions in which Puerto Rican populations are concentrated nor the heritage group differences in educational attainment, nativity, sex, or age distributions. At the same time, Latine heritage groups vary in numerous other ways which were not measurable with the present study’s data. Potentially relevant differences between Latine heritage groups are nonetheless discussed as possible explanations not examined in the study.

Subgroup differences in rates of substance use or substance use disorders represent one plausible contributor to differences in rates of overdose mortality. However, prior research has yielded mixed findings about which Latine heritage groups report the highest prevalence of substance use or substance use disorders. While several epidemiologic studies suggest that both the Puerto Rican and Mexican heritage groups have higher prevalence of drug use or drug use disorders than Cuban Americans [8–10], another national study identified South American and Cuban heritage as the groups with the highest prevalence of various types of drug use among several subgroups [11]. Previously-documented differences in *drug use* across Latine subgroups do not reach the extent of the gap in *drug overdose mortality* rates between Puerto Rican heritage and other Latine heritage groups, with overdose mortality rates in Puerto Ricans three times as high as any other group examined. Drug use prevalence estimates do not capture risk differences based on the types of drugs used, method of administration (e.g., injection), use patterns, trajectories, or risk practices, all of which shape risk of fatal overdose. For example, Puerto Ricans in the Northeast report use of heroin as well as cocaine [40], often substituted or contaminated with fentanyl [27, 41], heightening overdose vulnerabilities and providing a plausible explanation for the elevated overdose mortality rates of Puerto Ricans in the Northeast. Nonetheless, the rationales underlying the disparate rates of drug overdose deaths in Puerto Ricans in other US geographies with arguably weaker fentanyl footprints remain elusive and require additional research.

Although not examined in the present study, several social disadvantages may be disproportionately experienced by stateside Puerto Ricans across US geographies. For instance, some evidence suggests that Puerto Rican populations in the US are more residentially segregated than Mexican Americans, and that the link between residential segregation and poor health is more salient for Puerto Ricans than Mexican Americans [42–44]. The Puerto Rican population experiences the highest unemployment rate of all the Latine subgroups that were included in the present study, as well as the second-lowest median household income and lower rates of home ownership than Mexican, Cuban, or South American groups [19]. When considering exposure to trauma, which has been identified as a risk factor for overdose among individuals who use drugs [45–46], the Puerto Rican group has the highest percentage of veterans across all Latine heritage groups [19], and evidence from select US cities suggests that Adverse Childhood Experiences may also disproportionately impact Puerto Rican populations [47]. Moreover, rates of incarceration in the male US population ages 18-39 are higher in individuals of Puerto Rican heritage than any other Latine group [48]. Incarceration may foster risky drug use practices [49–50], elevate vulnerability to fatal overdose upon release [51], and deprive individuals of employment and housing opportunities that are critical to health and well-being; therefore, any disproportionate history of incarceration in the Puerto Rican population may potentially contribute to this group’s disparate rates of fatal overdose.

The Puerto Rican-heritage population in the US also represents the Latine subgroup with the highest prevalence of a disability, with 14.5% of the population aged 18-64 reporting a disability, compared to 7.5% among those of Mexican heritage [19], and research suggests that opioid overdose fatalities are overrepresented among nonelderly individuals with a disability, at least among Medicare enrollees [52]. The Puerto Rican population in the US is also the Latine subgroup with the highest risk for HIV infection via injection drug use [53–54]. Among individuals who use drugs, HIV infection is associated with increased risk of overdose, potentially via health factors and shared structural or behavioral risk factors [55]. Some researchers suggest that the at-birth US citizenship of Puerto Ricans enables individuals with a variety of health statuses [8], including individuals with HIV or HCV [56], to migrate from Puerto Rico to the mainland US, while immigration barriers for Latines from countries outside the US result in the migration of primarily healthier individuals from other Latine subgroups [57]. Foundational work by Deren and colleagues documented an “airbridge” [58–61] between Puerto Rico and the United States, wherein people who use or inject drugs and are vulnerable to HIV and overdose move across these geographies [60]. Due to this “airbridge,” some Puerto Ricans migrating to the US are more likely to need overdose prevention services than other Latine migrants. Moreover, as a colonial “territory” of the US, Puerto Rico is the closest entry point to the US for heroin and cocaine trafficking routes originating in South America [62]. Regardless of the cause, the disproportionately high rates of drug overdose mortality in Puerto Rican stateside populations underscore the importance of interventions and services that specifically target this population.

### 5.2 Association Between Nativity and Overdose Mortality Across Heritage Groups

The present study’s results indicate that the relationship between nativity and drug overdose mortality varies by Latine heritage group. Rates of drug overdose mortality were higher for the US-born than foreign-born in the Mexican, Cuban, Dominican, Central American, and South American heritage groups, yet the opposite was observed for the Puerto Rican heritage group. Consistent with prior research [25], drug overdose mortality rates were lower in US-born Puerto Ricans than Puerto Ricans not born in the 50 states/DC (i.e., those born in Puerto Rico). Although the often-referenced “immigrant paradox” predicts better health outcomes in immigrant populations than their US-born counterparts, some research has indicated that the immigrant paradox does not apply to all Latine subgroups nor all health-related measures [7, 9, 50]. A study using a nationally-representative sample of the US civilian, non-institutionalized adult population reported a higher lifetime prevalence of drug use among the US-born across all Latine heritage groups examined (Mexican, Puerto Rican, Cuban, Central American, and South American) [11]. Nonetheless, the present study’s results indicate that for the outcome of drug overdose mortality, the pattern of higher risk in the US-born may apply to Mexican, Cuban, Dominican, Central American, and South American groups but *not* the stateside Puerto Rican population.

Although we are unable to determine what portion of the Puerto Rico-born decedents in the present study had initiated injection drug use in Puerto Rico, a rich body of qualitative research has identified unique risk vulnerabilities among migrants to New York City who began injecting drugs while in Puerto Rico [40, 50, 63]. Cross-sectional studies, including various cycles of the National HIV Behavioral Surveillance (NHBS) study conducted by the CDC in New York City, suggest that Puerto Ricans who inject drugs and migrated from Puerto Rico are the most HIV-vulnerable ethnic group in New York City [40, 64]. Additionally, over the past 15 years, the NHBS has documented the sustained and sizeable (≥25%) presence of these migrants in New York City’s population of individuals injecting drugs [65].

Research also indicates that migrants’ HIV, HCV, and overdose risks began while injecting drugs in Puerto Rico, before US migration [40, 50, 63]. Often shaped by experiences in the carceral systems of Puerto Rico, individuals in this subpopulation report frequent syringe re-use and sharing after water rinsing and air-blowing, consistent speedball (cocaine and heroin) use, increased frequency of injection due to the experience of lower drug potency in the continental US compared to Puerto Rico, and practices of pooling money to purchase and share drugs and supplies [50, 63]. Many also express ambivalence toward medication-assisted treatment, due to ingrained messages from abstinence-only and religious-centered, shame-based treatment experiences in Puerto Rico [50]. Finally, negative law enforcement experiences and fear of arrest hinder this subpopulation from carrying syringes even though these are readily-available from syringe exchange programs in New York City [50, 63]. The LatCrit lens gains relevance when considering the service needs of these predominantly Spanish-monolingual migrants. Reductionist approaches to “Hispanic health,” which design health campaigns assuming that the Spanish language is uniform across Latin American cultures, are the norm, yet, judging by the overdose rates in the present study, are also likely ineffective.

To address the health vulnerabilities of migrants who initiated injection drug use in Puerto Rico, researchers have provided specific suggestions for harm reduction programs in US cities, including: hiring staff who speak and understand the Spanish spoken in Puerto Rico and training them in relevant community affiliations and Puerto Rico-specific drug lingo; providing services as near as possible to the locations where drugs are used daily; and capitalizing on the potential for collective action within the community of migrants who use drugs in order to bolster naloxone carrying [63]. The specificity of these recommendations illustrates the importance of approaching Latines not as one uniform group with a one-size-fits-all approach, but rather attending to the structural barriers, lived experiences, and cultural practices relevant to specific Latine subpopulations and communities. For example, rates of drug overdose mortality in the Mexican, Cuban, Dominican, Central American, and South American groups were higher among those born in the US. Hence, providing such US-born individuals with overdose prevention materials or services in Spanish may prove insufficient to effectively address their overdose risks. An anti-essentialist approach to Latine health acknowledges that not all Latine “speak Spanish, or want to” [16].

### 5.3 Missingness of Specific Hispanic Heritage

In the study’s examination of missingness of specific Hispanic heritage group, substantial state-level variation was observed; in New Mexico, for example, 86.50% of Hispanic overdose decedents were missing a specific Hispanic heritage group and were categorized under an ambiguous code corresponding to “Spanish” origin. It is unclear to what extent state-level variation in Hispanic group missingness reflects local differences in death certificate reporting procedures, as opposed to differences in the histories and identities of Hispanic populations across various regions. It is possible that members of Hispanic populations that have lived for numerous generations in states such as New Mexico or Colorado are more likely to be identified only with a panethnic term such as “Hispanic” or “Spanish,” in contrast to populations in states with more recent immigrants, where Hispanics may be more likely identified by a specific country or region of origin [66]. Aligned with LatCrit, these findings raise important questions about the preparedness (and rigor) of systems with the power to classify and categorize Hispanics (and any other racial/ethnic minority).

Additional research would be needed to determine ways to minimize missingness of Hispanic heritage group on death certificates and improve consistency between death certificate data and the way individuals self-identified during their lifetimes, bearing in mind both the limitations implicit in discrete categories of race/ethnicity and the sometimes-fluid self-identification of heritage throughout the life course [67]. Regardless, results from models with imputed Hispanic heritage suggest that using only cases without missing Hispanic heritage group may result in underestimates of drug overdose mortality rates for all Latine heritage groups, as observed rates were statistically significantly lower than the rates estimated with imputed data.

## 6. Limitations

Death certificate data provide only the most basic information about the decedent, restricting the study’s scope and the variables available for examination. Nativity was the only death certificate measure connected to cultural identification or language, yet nativity is a limited proxy for the complex and multidimensional process of acculturation. Both educational attainment and race/ethnicity are sometimes misreported on death certificates [30, 68]. In the present study, the imputation procedure addressed instances of *un*classified Hispanic heritage group but was unable to address any potential instances of *mis*classified Hispanic heritage in the mortality data. The multiple imputation procedure used in the present study also relied on assumptions about patterns of missingness as well as the presumption that every decedent identified as Hispanic “belonged to” one of the seven different Hispanic heritage groups.

The ethnicity categories in the data sources accommodated one heritage group only (even if an individual may have self-identified with multiple Hispanic heritage groups during life) and combined many diverse counties of origin into the categories of Central American and South American. Moreover, small numbers of deaths in subgroups, as well as limited racial categories in the mortality data, precluded disaggregation by race within Latine subgroups (e.g., Black or White Puerto Rican). Finally, the population estimates used to calculate rates are subject to sampling and non-sampling error [69].

## 7. Implications

The present study’s results add to the body of research underscoring the importance of health data disaggregation for the diverse Hispanic population [7–9, 13, 70–75]. For instance, when drug overdose mortality rates are presented for the Hispanic category overall, disproportionately high rates of overdose mortality in stateside Puerto Rican populations are obscured. Whenever feasible, disaggregating outcomes such as drug overdose mortality by specific Hispanic heritage can serve as a first step toward more fully addressing health risks for US Hispanics; this may also hold true for other health phenomena, including HIV [53], HCV [76], and COVID-19.

Disaggregating mortality data by nativity also has potential to inform more tailored interventions; however, results of the present study suggest that dividing the overall Hispanic group into only two nativity categories (US-born and foreign-born) may still mask substantial variation. For example, categorizing those born in Puerto Rico in a broad “US-born Hispanic” group would hide the unique linguistic and cultural needs of those born in Puerto Rico; at the same time, including those born in Puerto Rico in a broad “foreign-born Hispanic” category would entail placing the Puerto Rican-born, with elevated risk of overdose mortality relative to the US-born, in the same group as Mexican-born, Cuban-born, Central or South-American-born, with lower risk of overdose mortality relative to the US-born. As such, whenever possible, disaggregating by place of birth within specific Latine heritage groups (e.g., US-born Mexican, foreign-born Mexican) may be advisable, and when not possible to divide by heritage groups, expanding nativity to three categories for Latines (i.e., US-born [50 states/DC], territory-born [e.g., Puerto Rico], and foreign-born) may be another option for consideration. Admittedly, these categories are also limited in their portrayal of the multidimensional cultures they label. Nonetheless, they represent a move toward data disaggregation to more effectively promote population health.

## Data Availability

The data used in the study are restricted-access but are available via application from the National Center for Health Statistics.

## Acknowledgements

The authors wish to thank the National Center for Health Statistics for access to the mortality data, as well as the IPUMS for providing Census Bureau population data.

## Institutional Review Board

The Institutional Review Board of the University of Texas at San Antonio waived review of the present study’s analysis of the de-identified decedent data.

