## Appendix for "Variation in US Drug Overdose Mortality Within and Between Hispanic/Latine Subgroups: A Disaggregation of National Data"

### Specific Hispanic Heritage Identification on Death Certificates

On the U.S. Standard Death Certificate (2003 revision; https://www.cdc.gov/nchs/data/dvs/death11-03final-acc.pdf), five checkboxes are provided for the "decedent of Hispanic origin" question:

- "No, not Spanish/Hispanic/Latino;"
- "Yes, Mexican, Mexican American, Chicano;"
- "Yes, Puerto Rican;"
- "Yes, Cuban;"
- "Yes, other Spanish/Hispanic/Latino (Specify)" [with a blank line for a write-in response].

Ethnicity information on the death certificate is completed by a funeral director, who is instructed to request this information from the decedent's next of kin but may at times fill out this information based on observation instead (Arias et al., 2016). When mortality data is compiled by vital statistics, Hispanic origin is categorized according to a list of codes [https://www.cdc.gov/nchs/data/dvs/Appendix_D_Accessible_Hispanic_Origin_Code_List_Update_2011.pdf].

Per NCHS instructions, Hispanic heritage that is missing is coded as: Mexican if the place of birth listed on the death certificate is Mexico; Puerto Rican heritage if the place of birth is Puerto Rico; and Cuban heritage if the place of birth is Cuba; other places of birth do not result in recoding of missing heritage [https://www.cdc.gov/nchs/data/dvs/2014_PT11_NOV2014.pdf].

The National Center for Health Statistics has conducted several evaluations of race and Hispanic origin reporting on death certificates, most recently using 1999-2011 linked death certificate race/ethnicity responses with self-reported race/ethnicity responses from the Current Population Survey (Arias et al., 2016). In this NCHS report, the Central and South American subgroups evidenced higher levels of misclassification than the Mexican, Puerto Rican, or Cuban subgroups, yet the classification ratios for specific Hispanic heritage groups differed over the three different time periods examined and also varied by nativity and the region, rurality, and co-ethnic concentration of the decedent's area of residence (Arias et al., 2016).

Although providing very detailed and comprehensive information about the misclassification of Hispanic heritage for deaths of all causes, the results from the NCHS report could not be directly applied to the present study in particular. While our analysis is focused on missingness of specific heritage, the NCHS report combines *both* missingness and misclassification of heritage—that is, including individuals for whom specific Hispanic heritage was missing as well as individuals for whom heritage was not missing but was instead inaccurate (for example, someone who self-identified as Central or South American during life but was recorded on the death certificate as Mexican or even as Non-Hispanic). Furthermore, in the NCHS report, the "other Hispanic" group encompasses individuals identified as Dominican and Spaniard as well as those for whom Hispanic heritage was missing, while our study examines Dominican heritage, Spaniard, and missing heritage as distinct groups.

### Approaches for Other Types of Missing Data on Death Certificates

Due to a lack of published studies directly addressing missing Hispanic heritage on drug overdose death certificates, we examined approaches used to handle other types of missing data on death certificates.

#### Missing data on drug type

In drug overdose mortality research, one of the most prominent types of missing data is the type of drug involved in overdose (Boslett et al., 2019; Slavova et al., 2015), which leads to underestimates for mortality rates of overdoses involving specific drug types (e.g., heroin, opioids; Ruhm, 2018). In 2017, drug type was missing for approximately 22% of overdose deaths at the national level, yet missingness varied widely, ranging from 2% missing in one county to nearly 95% missing in another county within the same state (Jones et al., 2019). Several different approaches to missingness of drug type have been identified in the literature:

1. CDC reports (Mortality and Morbidity Weekly Reports and Data Briefs) do not generally use imputation/correction for drug missingness; however, CDC only presents state-level drug-specific overdose mortality rates (e.g., opioid-related overdose mortality rates) for those states with less than 20% missingness of drug type (i.e., meeting "good" or "very good to excellent" reporting standards, which are also based on levels of change in missingness across years; Rudd et al., 2016).

2. In at least one study, researchers assigned opioid involvement to overdose deaths with missing drug type by assuming the same percentage of opioid involvement observed for the drug overdose deaths that were not missing drug type (Buchanich et al., 2018). This approach was examined in an analysis of data from one county in Indiana, in which death certificate records were linked to toxicology reports to compare imputed and actual data on opioid involvement; results indicated that this imputation approach underestimated opioid involvement minimally (Gupta et al., 2020).

3. Multiple Imputation: Ruhm (2018) used multiple imputation for missing opioid involvement on the death certificate, in a model with decedent demographics and county characteristics. Boslett et al. (2020) assessed the predictive accuracy of results from Ruhm's methodology, as well as additional imputation models; results indicated that imputation via either logistic regression or random forest models provided similar performance, with predicted accuracy improved by including contributing causes of death in the models. Gupta et al. (2020) also replicated Ruhm's type of imputation model, using linked toxicology reports in one Indiana county, finding that the imputation model overestimated opioid involvement minimally.

#### Specific Asian race

Thompson et al. (2018) used multiple imputation to assign specific Asian groups (e.g., Korean and Vietnamese) to decedents from states in which these groups were classified only as "other Asian" prior to the implementation of updated race reporting standards for mortality data.

### Imputation Approach in the Present Study

#### Missingness of Hispanic Heritage in the Present Study

The ethnicity information for 5,095 of the 29,137 (17.48%) Hispanic drug overdose decedents in the present study lacked sufficient detail to classify these decedents into one of the seven Hispanic heritage groups (Mexican, Puerto Rican, Cuban, Dominican, Central American, South American, or Spaniard). This group of 5,095 decedents could possibly include some individuals who identified as "Hispanic" but did not identify with any of the seven heritage groups. For the purposes of the present study, however, we consider only seven plausible Hispanic heritage groups based on Census categories (Ennis et al., 2011).

The ethnicity codes classified as "missing Hispanic heritage" in the present study are listed below, ordered based on descending frequency:

- Code 282, for "Spanish," comprised 38.65% of the missing Hispanic heritage group.
- Code 281, for "Hispanic," comprised 31.15% of the missing Hispanic heritage group.
- Code 280, for "Other Spanish Checkbox," comprised 14.01% of the missing Hispanic heritage group.
- Code 299, for "Other Spanish," comprised 10.17% of the missing Hispanic heritage group.
- Codes 250-259, for "Latin American," comprised 4.36% of the missing Hispanic heritage group.
- Code 220, for "Central and South America," comprised 1.08% of the missing Hispanic heritage group. This code was included under missing specific Hispanic heritage in the present study because it lacks specificity to determine whether the decedent should be classified as Central American or as South American.
- Code 286, for "Spanish American," comprised 0.51% of the missing Hispanic heritage group.
- Code 283, for "Californio," comprised 0.04% of the missing Hispanic heritage group.
- Code 287, for "Spanish American Indian," comprised 0.02% of the missing Hispanic heritage group.
- Code 289, for "Mestizo," comprised 0.02% of the missing Hispanic heritage group.

As observed in the preceding list, the majority of the missing Hispanic heritage group corresponded to codes that could plausibly apply to many different Hispanic heritage groups, such as code 281, "Hispanic," as opposed to a code such as code 283, "Californio" (0.04%), which would not likely apply to all heritage groups, or the code 220 (1.08%), which would apply only to Central or South American heritage.

Supplemental Table 1 provides the state-level percentages of missing Hispanic heritage group that corresponded to each code for the Hispanic drug overdose decedents with missing heritage in the present study. In accordance with CDC confidentially guidelines for subnational data, we suppress all state-level percentages based on fewer than 10 deaths. Cells are shaded according to value, with the *darkest* cells indicating the highest percentages for each state. As presented in Supplemental Table 1, the codes most frequently observed for the "Hispanic heritage missing" group varied across states. For example, the most frequently observed code for Hispanic heritage missing was code 299 ("Other Spanish") in New York (81.29% of missing), code 281 ("Hispanic") in Texas (89.49% of missing), and code 280 ("Other Spanish Checkbox") in Virginia (90.00% of missing).

### Supplemental Table 1

*Percentages of the "Hispanic heritage missing" group corresponding to each ethnicity code, by state, for drug overdose deaths among Hispanics, 2015-2019*

| code | 220 Central and South American | 250 Latin American | 280 Other Spanish Checkbox | 281 Hispanic | 282 Spanish | 286 Spanish American | 299 Other Spanish |
|---|---|---|---|---|---|---|---|
| AZ | - | - | 23.23 | 34.34 | 39.39 | - | - |
| CA | - | 11.10 | 6.68 | 63.00 | 18.29 | - | - |
| CO | - | 2.45 | 4.90 | 36.60 | 52.61 | 3.10 | - |
| CT | - | - | 31.25 | 43.75 | - | - | - |
| FL | 41.60 | - | 26.40 | 12.80 | 14.40 | - | - |
| GA | - | - | - | 43.48 | - | - | - |
| HI | - | - | 68.97 | - | - | - | - |
| IN | - | - | - | 65.52 | - | - | - |
| LA | - | - | - | 63.64 | - | - | - |
| MA | - | - | 70.25 | 27.85 | - | - | - |
| MD | - | - | 52.63 | - | - | - | - |
| MI | - | - | 74.03 | 16.88 | 7.14 | - | - |
| NJ | - | 12.36 | 32.58 | 31.46 | 17.98 | - | - |
| NM | - | - | - | - | 98.50 | - | - |
| NV | - | - | - | - | 58.33 | - | - |
| NY | - | - | 8.87 | 5.32 | 4.19 | - | 81.29 |
| OK | - | - | - | 72.73 | - | - | - |
| PA | - | 14.46 | 32.53 | 37.35 | 12.05 | - | - |
| TN | - | - | 60.00 | - | - | - | - |
| TX | - | - | - | 89.49 | 5.41 | - | - |
| UT | - | - | - | 50.65 | 33.77 | - | - |
| VA | - | - | 90.00 | - | - | - | - |
| WA | - | - | 14.08 | 52.11 | 23.94 | - | - |

*Notes.* Percentages based on fewer than 10 deaths are suppressed ("-") per CDC confidentiality guidelines. States and categories with only suppressed values are not presented in the table. Percentages may not sum to 100 across rows due to rounding and suppressed values.

### Associations Between Missingness and Observed Variables

In order to examine associations between "missingness of specific Hispanic heritage" and other variables available via the death certificate, a binary variable was created for missingness of Hispanic heritage. Missingness was independently regressed on each of several variables, including decedent demographics and characteristics of the deaths, as well as the percentage of the Hispanic population identified as Mexican, Puerto Rican, Cuban, Dominican, Central American, or South American in the county or state of residence of the decedent, per 2015-2019 American Community Survey five-year estimates via IPUMS. (State of residence was used when detailed Hispanic heritage population counts were not available for the county of residence.) All variables were binary (yes=1; no=0), with the exception of continuous measures for age and percent Mexican/Puerto Rican, etc. Odds ratios and accompanying 95% Confidence Intervals are provided in Supplemental Table 2.

### Supplemental Table 2

*Results of binomial logistic regressions of missingness of specific Hispanic heritage on each characteristic*

| Characteristic | Odds Ratio (95% CI) | p value |
|---|---|---|
| Age | 1.007 (1.004-1.009) | <0.001 |
| **Sex** | | |
| Female | 1.392 (1.301-1.489) | <0.001 |
| **Drug involved in the overdose** | | |
| Heroin | 1.112 (1.040-1.188) | 0.002 |
| Natural and semi-synthetic opioid | 1.617 (1.498-1.745) | <0.001 |
| Cocaine | 0.680 (0.629-0.734) | <0.001 |
| Psychostimulant with abuse potential[a] | 1.538 (1.434-1.650) | <0.001 |
| Benzodiazepine | 1.280 (1.176-1.392) | <0.001 |
| Alcohol | 0.937 (0.856-1.026) | 0.160 |
| Synthetic opioid | 0.665 (0.623-0.710) | <0.001 |
| **Race** | | |
| White | 0.712 (0.624-0.812) | <0.001 |
| Black | 1.118 (0.934-1.338) | 0.226 |
| Other | 1.838 (1.520-2.221) | <0.001 |
| **Educational attainment** | | |
| 8th or less | 0.390 (0.338-0.449) | <0.001 |
| 9-12th, no diploma | 0.943 (0.876-1.015) | 0.118 |
| High School/GED | 1.026 (0.966-1.090) | 0.406 |
| Some College | 1.162 (1.066-1.267) | 0.001 |
| Associate's/Bachelor's | 1.310 (1.180-1.455) | <0.001 |
| Graduate | 1.309 (0.999-1.716) | 0.051 |
| Unknown | 2.068 (1.762-2.426) | <0.001 |
| **Status** | | |
| Single | 0.895 (0.842-0.952) | <0.001 |
| Married | 0.890 (0.823-0.962) | 0.003 |
| Divorced | 1.144 (1.057-1.239) | 0.001 |
| Widow | 1.177 (0.976-1.420) | 0.089 |
| Unknown | 2.964 (2.438-3.602) | <0.001 |
| **Intent** | | |
| Unintentional | 0.931 (0.835-1.038) | 0.201 |
| Intentional | 1.184 (1.044-1.343) | 0.008 |
| Undetermined | 0.841 (0.685-1.033) | 0.099 |
| **Nativity** | | |
| Born in 50 states/DC | 5.237 (4.735-5.791) | <0.001 |
| Born in a US territory[b] | 0.003 (0.001-0.011) | <0.001 |
| Foreign-born[b] | 0.360 (0.325-0.399) | <0.001 |
| **Method of disposition** | | |
| Burial | 0.678 (0.636-0.724) | <0.001 |
| Cremation | 1.689 (1.585-1.800) | <0.001 |
| **US Census region** | | |
| Northeast | 0.628 (0.583-0.676) | <0.001 |
| Midwest | 0.449 (0.394-0.511) | <0.001 |
| South | 0.386 (0.354-0.420) | <0.001 |
| West | 3.357 (3.154-3.574) | <0.001 |
| **Hispanic population composition in decedent's county/state of residence[c]** | | |
| Percent Mexican | 1.013 (1.012-1.014) | <0.001 |
| Percent Puerto Rican | 0.976 (0.974-0.978) | <0.001 |
| Percent Cuban | 0.906 (0.896-0.916) | <0.001 |
| Percent Dominican | 0.993 (0.990-0.996) | <0.001 |
| Percent Central American | 0.959 (0.955-0.964) | <0.001 |
| Percent South American | 0.945 (0.940-0.950) | <0.001 |

*Notes.* All predictor variables (except age and percent heritage) are binary variables (1=yes; 0=no). [a]Category excludes cocaine [b]Decedents with recorded place of birth in Mexico, Cuba, or Puerto Rico are not missing heritage due to coding procedures of NCHS which involve assigning heritage, whenever missing, based on place of birth in Mexico, Cuba, or Puerto Rico. [c]Based on 2015-2019 American Community Survey five-year estimates for the county of residence of decedent, or for the state of residence when population estimates by Hispanic heritage were not available for the particular county of residence.
Data from 2015-2019 for 29,137 drug overdose deaths in which the decedent was identified as of Hispanic heritage.

### Assumptions for the Imputation Model

The associations between missingness of specific Hispanic heritage and observed variables in the data supported *Missing At Random* (MAR) rather than *Missing Completely At Random* (MCAR), although the possibility of *Missing Not At Random* (MNAR) cannot be discounted (Mack et al., 2018).

If missingness depends on the values of the missing data themselves, or other unobserved variables (MNAR), this would represent a concern for the use of a multiple imputation model. Although the National Center for Health Statistics' report on misclassification of Hispanic heritage suggests that Central and South American decedents are more often *misclassified* (Arias et al., 2016) in overall mortality data, it is unclear whether these groups are more likely to be *missing* specific Hispanic heritage in mortality data as opposed to being incorrectly classified in mortality data, nor whether any difference would persist after accounting for observed variables available.

One plausible concern is that perhaps Hispanic heritage groups without a designated checkbox on the death certificate (that is, Central American, South American, Dominican, or Spaniard) would be more likely to be recorded without a specific heritage group due to the need for the funeral director to write out a response to heritage group rather than only checking a box. However, US Census Questionnaires use a similar Hispanic origin question (with a checkbox for Mexican, Puerto Rican, or Cuban heritage and a line for a write-in response next to the checkbox for "other Hispanic heritage"), yet on Census questionnaires even individuals who identify as Mexican in a separate ancestry question sometimes bypass the Mexican checkbox in the Hispanic origin question and write in a generic term such as "Hispanic" or "Latino" next to the "Other Hispanic" checkbox (Alba, 2007; Martin, 2006; Ramirez, 2005; Suro, 2002). This suggests that a generic response of "Hispanic," "Latino," or "Other Hispanic" may not necessarily only be used for individuals of heritage groups without a designated checkbox. In addition, in the case of reporting on the death certificate, a generic response such as "Hispanic" may be used when the funeral director is unsure of a more specific heritage beyond Hispanic, and this situation may apply whether or not the decedent's heritage category has a designated checkbox on the death certificate.

Based on the preceding considerations, we use multiple imputation under the *MAR* assumption.

### Imputation Procedure in the Present Study

A polytomous logistic regression imputation model was used for multiple imputation of Hispanic heritage group, with 20 imputed datasets. Analyses were conducted in Stata/MP 16.1 via *mi impute mlogit*. Variables included in the model comprised decedent demographics, characteristics of the death, and county/state-level percentages of Hispanics identified as Mexican, Puerto Rican, Cuban, Dominican, Central American, or South American, respectively (per IPUMS American Community Survey five-year 2015-2019 estimates). All categorical variables were recoded into binary measures for each category. Supplemental Table 3 provides details about each variable in the model.

### Supplemental Table 3

*Description of variables included in imputation model*

| Characteristic | Variable type | Notes |
|---|---|---|
| **Individual-level data from death certificate** | | |
| Age | Continuous | |
| **Sex** | | |
| Female | Yes/no | |
| **Drug involved in overdose** | | |
| Heroin | Yes/no | “multiple cause of death” with ICD-10 code T40.1 |
| Natural/semi-synthetic opioid | Yes/no | “multiple cause of death” with ICD-10 code T40.2 |
| Cocaine | Yes/no | “multiple cause of death” with ICD-10 code T40.5 |
| Psychostimulant[a] | Yes/no | “multiple cause of death” with ICD-10 code T43.6 |
| Benzodiazepine | Yes/no | “multiple cause of death” with ICD-10 code T42.4 |
| Alcohol | Yes/no | “multiple cause of death” with ICD-10 code T51.0 |
| Synthetic opioid excl. methadone | Yes/no | “multiple cause of death” with ICD-10 code T40.4 |
| **Race** | | |
| White | Yes/no | |
| Black | | ommitted |
| Other | Yes/no | |
| **Educational attainment** | | |
| 8th or less | Yes/no | |
| 9-12th, no diploma | Yes/no | |
| High School/GED | Yes/no | |
| Some College | Yes/no | |
| Associate’s/Bachelor’s | Yes/no | |
| Graduate | Yes/no | |
| Unknown | | ommitted |
| **Status** | | |
| Single | Yes/no | |
| Married | Yes/no | |
| Divorced | Yes/no | |
| Widowed | | ommitted |
| Unknown | Yes/no | |
| **Intent** | | |
| Unintentional | Yes/no | |
| Intentional | Yes/no | |
| Undetermined | | ommitted |
| **Nativity** | | |
| Born in 50 states/DC | Yes/no | |
| Born in a US territory[b] | Yes/no | |
| Foreign-born[b] | | ommitted |
| **Method of disposition** | | Other methods of disposition ommitted |
| Burial | Yes/no | |
| Cremation | Yes/no | |
| **US Census region** | | |
| Northeast | Yes/no | |
| Midwest | Yes/no | |
| South | Yes/no | |
| West | | ommitted |
| **County/State Data from IPUMS American Community Survey 2015-2019 Five Year Estimates** | | |
| **Hispanic population composition in decedent’s county/state of residence[c]** | | |
| Percent Mexican | Continuous | Percent _______ out of total Mexican, Puerto Rican, Cuban, Dominican, Central or South American population in county or state of residence |
| Percent Puerto Rican | Continuous | |
| Percent Cuban | Continuous | |
| Percent Dominican | Continuous | |
| Percent Central American | Continuous | |
| Percent South American | Continuous | |

*Notes.* [a]Category excludes cocaine [b]Decedents with recorded place of birth in Mexico, Cuba, or Puerto Rico are not missing heritage due to coding procedure of NCHS. [c]Based on 2015-2019 American Community Survey five-year estimates for the county of residence of decedent, or for the state of residence of the decedent when population estimates by Hispanic heritage were not available for the county of residence of the decedent.
